# Parental acceptance of and preferences for administration of routine varicella vaccination in the UK: a study to inform policy

**DOI:** 10.1101/2022.07.05.22277268

**Authors:** Susan M. Sherman, Nicola Lingley-Heath, Jasmine Lai, Julius Sim, Helen Bedford

## Abstract

**Objectives:** To explore acceptability of and preferences for the introduction of varicella vaccination to the UK childhood immunisation schedule.

**Design:** We conducted an online cross-sectional survey exploring parental attitudes towards vaccines in general, and varicella vaccine specifically, and their preferences for how the vaccine should be administered.

**Participants:** 596 parents (76.3% female, 23.3% male, mean age 33.4 years) whose youngest child was aged 0-5 years.

**Main outcome measures:** Willingness to accept the vaccine for their child and preferences for how the vaccine should be administered (in combination with the MMR vaccine [MMRV], on the same day as the MMR vaccine but as a separate injection [MMR+V], on a separate additional visit).

**Results:** 74.0% of parents (95% CI 70.2% to 77.5%) were extremely/somewhat likely to accept a varicella vaccine for their child if one became available, 18.3% (95% CI 15.3% to 21.8%) were extremely/somewhat unlikely to accept it and 7.7% (95% CI 5.7% to 10.2%) were neither likely nor unlikely. Reasons provided by parents likely to accept the vaccine included protection from complications of chickenpox, trust in the vaccine/healthcare professionals, and wanting their child to avoid their personal experience of chickenpox. Reasons provided by parents who were unlikely included chickenpox not being a serious illness, concern about side effects, and believing it is preferable to catch chickenpox as a child rather than as an adult. A combined MMRV vaccination or additional visit to the surgery were preferred over an additional injection at the same visit.

**Conclusions:** Most parents would accept a varicella vaccination. These findings highlight parents’ preferences for varicella vaccine administration, information needed to inform vaccine policy and practice and development of a communication strategy.

## Introduction

### Varicella Infection

Varicella (chickenpox) is a common, highly infectious disease caused by the varicella-zoster virus (VZV) with a peak age for infection in the UK before the age of five years.[1] Although chickenpox is generally mild, it can be severe in immunocompromised individuals, adults, pregnant women and neonates. However, most children requiring hospitalisation for complications are previously healthy.[2] Following a primary infection with VZV the virus remains dormant in the dorsal root ganglia and can reactivate, causing herpes zoster (shingles). This typically occurs later in life due to reduced immunity, but it can also affect younger people.

Although varicella is not a notifiable disease in England and Wales,[3] a recent study estimated that there were, on average, 4,694 hospital admissions with varicella per year in England between 2004 and 2017, of which 38.14% had known complications such as skin, neurological or gastrointestinal problems.[4] Admissions were highest in the 0-1 years age category, while complications were highest in adult age groups; they were also higher in children over 1 year of age compared to younger children. Hobbelen et al estimate that the cost of varicella to the NHS for 2013/14 was £6.8 million.[5]

### Varicella Vaccine

Safe and effective live attenuated varicella vaccines have been available since the 1980s, with the reported effectiveness of one dose about 80% against all varicella and almost 100% against moderate to severe varicella disease.[6] The World Health Organization (WHO) recommends that the vaccine should be included in routine vaccination programmes in countries where the infection is an important public health burden. As of 2018, 36, mainly higher-income, countries had implemented a universal varicella vaccination programme. Varicella vaccine can be given to individuals over one year of age with at least a four-week interval between the two doses and administered as a monovalent vaccine (V) or as a quadrivalent vaccine combined with measles, mumps and rubella (MMRV). The monovalent and quadrivalent vaccines have both been demonstrated to be safe and effective, although there is a small increased risk of fever and febrile seizures associated with MMRV vaccine given to children aged two or younger compared with MMR + V.[7] Nevertheless, several countries such as Germany, Australia and the US routinely offer MMRV.

A range of approaches can be taken in implementing the two-dose varicella vaccination within the routine childhood vaccination schedule. The most common schedule is a first dose at 12–18 months with a second between 4-6 years, although this can be administered using a shorter interval, with at least 3 months between first and second dose.[8] In the UK, if a monovalent vaccine were offered at 12 months at the same time as existing vaccines this would require an additional injection, making five in total.

### UK varicella vaccination policy

Currently in the UK, a selective vaccination policy recommends two doses of a single antigen varicella vaccine for family members of immunocompromised individuals and healthcare workers.[9] Varicella vaccine is also available on a private basis, although it is not possible to estimate coverage.

In the UK in 2010, the Joint Committee on Vaccination and Immunisation (JCVI) recommended that a two-dose childhood varicella vaccination programme should not be introduced, on the basis that it would not be cost-effective. Subsequently, consideration was given to introducing a one dose programme but modelling showed this too would not be cost-effective.[10] In considering whether to introduce a varicella vaccine in the UK, key issues examined by the JCVI relate to concerns that by introducing a vaccination programme there would be reduced exposure to VZV, which is thought to play a role in boosting immunity among immune individuals. Hypothetically, without this boosting, there could be an increase in the incidence of herpes zoster, making the programme not cost-effective. A herpes zoster vaccine programme for adults aged 70 to 79 was introduced in 2013, which may lessen this concern. It is also necessary to ensure very high varicella vaccine uptake rates, since less than optimal uptake could reduce but not eliminate infection and increase the age of acquisition of varicella to older age groups, in whom disease is more severe.[11]

The introduction of universal varicella vaccine in the UK is being re-considered.[4] Ensuring the successful introduction of any childhood vaccine programme requires an understanding of parents’ knowledge and attitudes regarding the disease and acceptability of the vaccine to inform policy and development of a vaccine communication strategy. However, UK research on current parental attitudes to childhood varicella and varicella vaccine is limited. In one 2007 study of over 800 parents in three primary care trusts, most rated the severity of chickenpox to be no more than unpleasant with only 40% saying they would accept a vaccine if it were available.[12] In contrast, 61% parents attending two London hospitals considered varicella to be serious, with 67% accepting a vaccine,[13] and more recently, in a survey of 1,510 parents commissioned by Public Health England (PHE) in early 2020, 66% of parents would accept a varicella vaccine. Parents in this survey considered chickenpox to be the least serious (44% serious or fairly serious) compared with other vaccine-preventable childhood infections (L Letley, personal communication, 2022).

This study aimed to explore parents’ knowledge of and views regarding varicella infection and varicella vaccination in the UK, including preferences for administration (combination or separate vaccine) and timing of vaccination, with the findings intended to inform any future implementation of routine varicella vaccination.

## Method

### Design

We conducted a cross-sectional survey between July and August 2021. Participants completed the survey online on the web-based survey tool “Qualtrics”.

### Participants

Participants (n=601) were recruited through Prolific (https://prolific.co/), an online research panel. Participants were eligible to take part if they were resident in the UK and their youngest child was born between 2017 and 2021. Of 628 people who began the survey, 605 completed it (96.3% completion rate). Three participants were omitted from the sample as they did not meet quality control checks and one was omitted as they left their country of residence blank.

Participants were paid £1.75 per completed survey.

### Sample size

A sample size of 601 was chosen to provide appropriately precise estimates of proportions, with a margin of error no worse than ±4%, at a 95% confidence level.

### Measures

The questionnaire was developed based on a literature review of determinants of childhood vaccine uptake. The final questionnaire is available online (https://osf.io/nxtuk/?view_only=0002498feee44988bc7d763c1ba34d1c [peer review link to be updated on acceptance with public link]).

Participants were asked for demographic details of themselves and their child(ren). Questions were asked about: chickenpox and parental perceptions of its risk and seriousness; chickenpox vaccination and preferences for how it should be administered; and attitudes towards vaccination generally and towards sources of vaccination information. Participants were asked to respond with their youngest child (‘index child’) in mind.

Specifically, we asked participants to report their age, gender, ethnicity, religion, highest level of education or professional qualification, their current working situation, total household income, marital status and whether they had a disability. We also asked for the age and gender of their (up to four) youngest child(ren) and whether they had heard of chickenpox prior to this survey,

All participants who had previously heard of chickenpox were asked whether their child had already had chickenpox. We then asked participants to indicate on a 5-point Likert scale (from strongly agree to strongly disagree) the extent to which they were worried that their child might get chickenpox, and if a friend’s child had chickenpox, the extent to which they would try to make sure their child had close contact with them in the hope they would catch chickenpox (both questions to parents whose child had not already had it). We asked all parents to indicate their agreement with statements: “chickenpox is usually a mild disease in healthy children”; “chickenpox can cause serious complications”; “it is better to have chickenpox when you are a child than when you are an adult”.

We then asked participants if their child had already had the chickenpox vaccine and, if they had not, we informed them that it is already recommended for children in some countries and asked them to rate on a 5-point scale how likely it was (from extremely unlikely to extremely likely) that they would accept a vaccine to protect against chickenpox for their child if one was recommended in the UK; we then asked those parents if *any* of their children had had chickenpox.

We then provided *all* parents (including those whose child had already had the vaccine and those who had not previously heard of chickenpox) with some basic information about the infection and the vaccine and informed them that the chickenpox vaccine is already recommended for children in some countries (see Appendix). They were asked to rate on a 5-point scale how likely it was (from extremely unlikely to extremely likely) that they would accept a vaccine to protect against chickenpox for their child if one was recommended in the UK. We asked a follow-up open-ended question asking for the main reason why they would be likely or unlikely to accept a chickenpox vaccine for their child.

Next, we explored participants’ preferences for administration of varicella vaccine. Participants were provided with three possible scenarios for each of two vaccine doses for how varicella vaccine might be included in the UK vaccination schedule, and they were asked to indicate for each scenario how likely they would be to have the vaccination for their child. The three scenarios were: 1) chickenpox vaccine given as a combination 4-in-1 vaccine with MMR (MMRV) at 12 months (at 3 years 4 months for the 2^nd^ dose); 2) given as a separate vaccine at the same time as the vaccines given at 12 months (at the same time as the pre-school MMR vaccine for the second dose); 3) given separately at another visit after the vaccines given at 12 months (at 15-18 months for the 2^nd^ dose). Parents were also asked the maximum number of injections they considered acceptable for their child on any one occasion.

To explore parental attitudes to vaccination in general, we modified the WHO SAGE Vaccine Hesitancy Scale (VHS) [14,15] for use in a UK setting. Although developed to measure vaccine hesitancy among parents, some of the statements were adapted for this study to assess general attitudes as opposed to hesitancy specifically. Perceptions of vaccine safety and effectiveness were sought by including definitions of ‘safe’ (‘means serious side effects are rare’) and ‘effective’ (‘means that most vaccines give good protection’) to ensure consistent interpretation by respondents. The scale is composed of five-point Likert items (scored 1–5). Two subscale scores were derived by summing scores on these individual items: a ‘lack of confidence score’ (from eight items; possible range 5–40) and a ‘risk’ score (from two items; possible range 2–10) (see Table 4). In each case higher scores indicate greater hesitancy.

Participants were asked to identify their sources of information about vaccination (all sources and then main source) from a list of 19 options based on those provided in the PHE immunisation tracking surveys conducted among parents of young children.[16] Lastly, they were asked to indicate the extent to which they agreed or disagreed with a series of statements about trust in various sources of vaccination advice.

Three unrelated attention check questions were included as a quality control measure. Participants were excluded if they answered two or more of these questions incorrectly.

The study was reviewed and approved by Keele University’s Psychology Research Ethics Committee (reference: PS-210200).

### Analysis

Descriptive statistics (means, standard deviations, counts and percentages) are provided for all measures. The open-ended responses for reasons why participants were likely or unlikely to have a chickenpox vaccination, were analysed with content analysis using an emergent coding approach, whereby codes were identified from the data.[17]

Potential predictors of vaccination likelihood were analysed through ordinal logistic regression. Predictors were identified *a priori* and included: parental demographics (age and gender); whether the respondent had ever refused a vaccine for his or her child; whether the index child had had chickenpox; a belief that chickenpox is usually a mild disease in healthy children; a belief that chickenpox can cause serious complications; and the hesitancy subscales (lack of confidence and risk). The proportional odds assumption of the analysis was checked and statistical significance was set at *p* ≤ .05 (two-tailed). Estimates are presented as odds ratios, with 95% confidence intervals (CIs). The predictive power of the whole regression model is expressed by the Nagelkerke pseudo-*R*^2^ statistic, which ranges from 0 to 1, with higher values indicating greater predictive power. The predictive power of an ordinal variable (which is represented by more than one odds ratio) is expressed by the increase in the Nagelkerke *R*^2^ that its addition to the model produced.

## Results

The survey was completed by a total of 601 participants. Five had not previously heard of chickenpox and so their data were not analysed further leaving 596 participants aged between 19 and 50 years (mean=33.4, standard deviation=5.2) who were included in the data analysis. Participant characteristics and those of their children are detailed in Table 1.

**Table 1.**
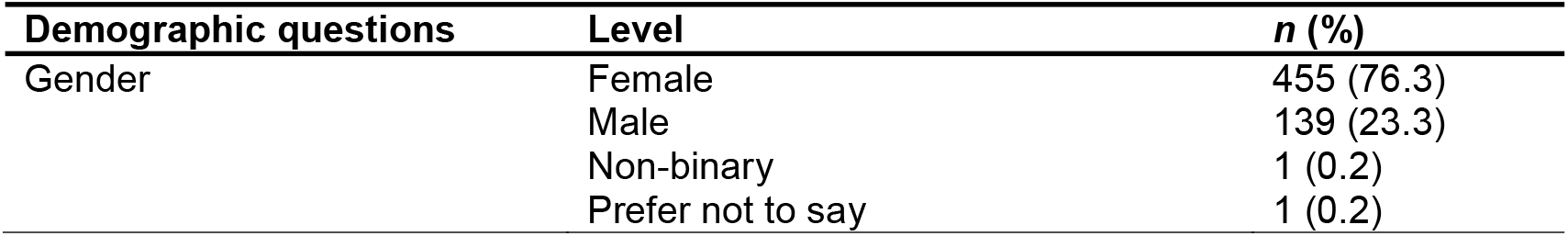
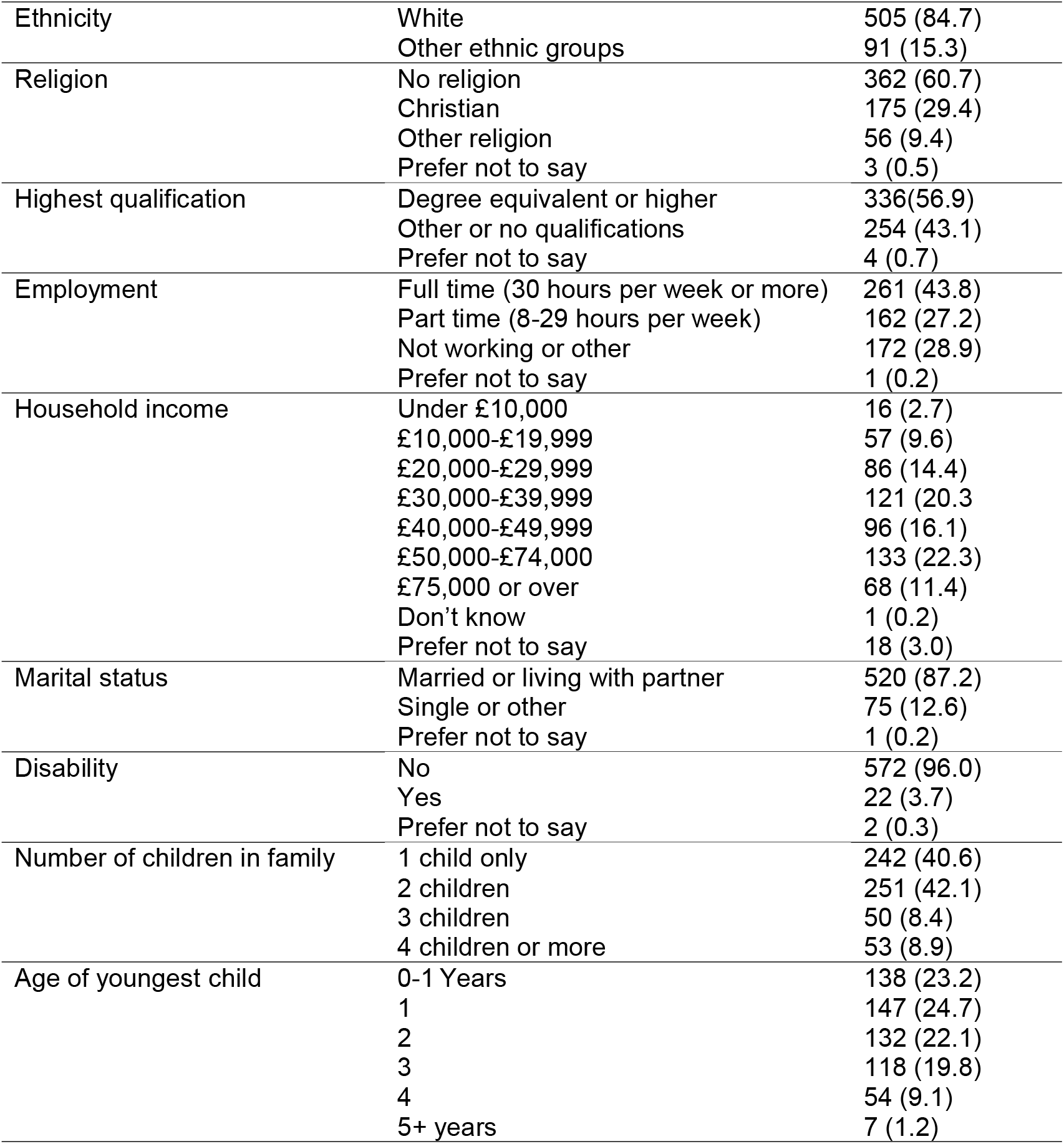
Participant characteristics

### Parents’ experiences of and attitudes to varicella infection

Parents generally considered varicella to be mild in children but more severe in adulthood. However, over half agreed that it could cause serious complications (Table 2). Over half the parents disagreed with intentionally infecting their child with varicella through contact with other infected children, and those whose child had not already had chickenpox expressed worry that their child would catch it. Vaccination intention is shown in Table 3; of 546 participants, after receiving information about varicella and the vaccination, 404 (73.9%; 95% CI 70.2% to 77.5%) were extremely/somewhat likely to accept a chickenpox vaccine for their child if one became available, 100 participants (18.2%; 95% CI 15.3% to 21.8%) were extremely/somewhat unlikely to accept it, and 42 participants (7.7%; 95% CI 5.7% to 10.2%) were neither likely nor unlikely.

**Table 2.**
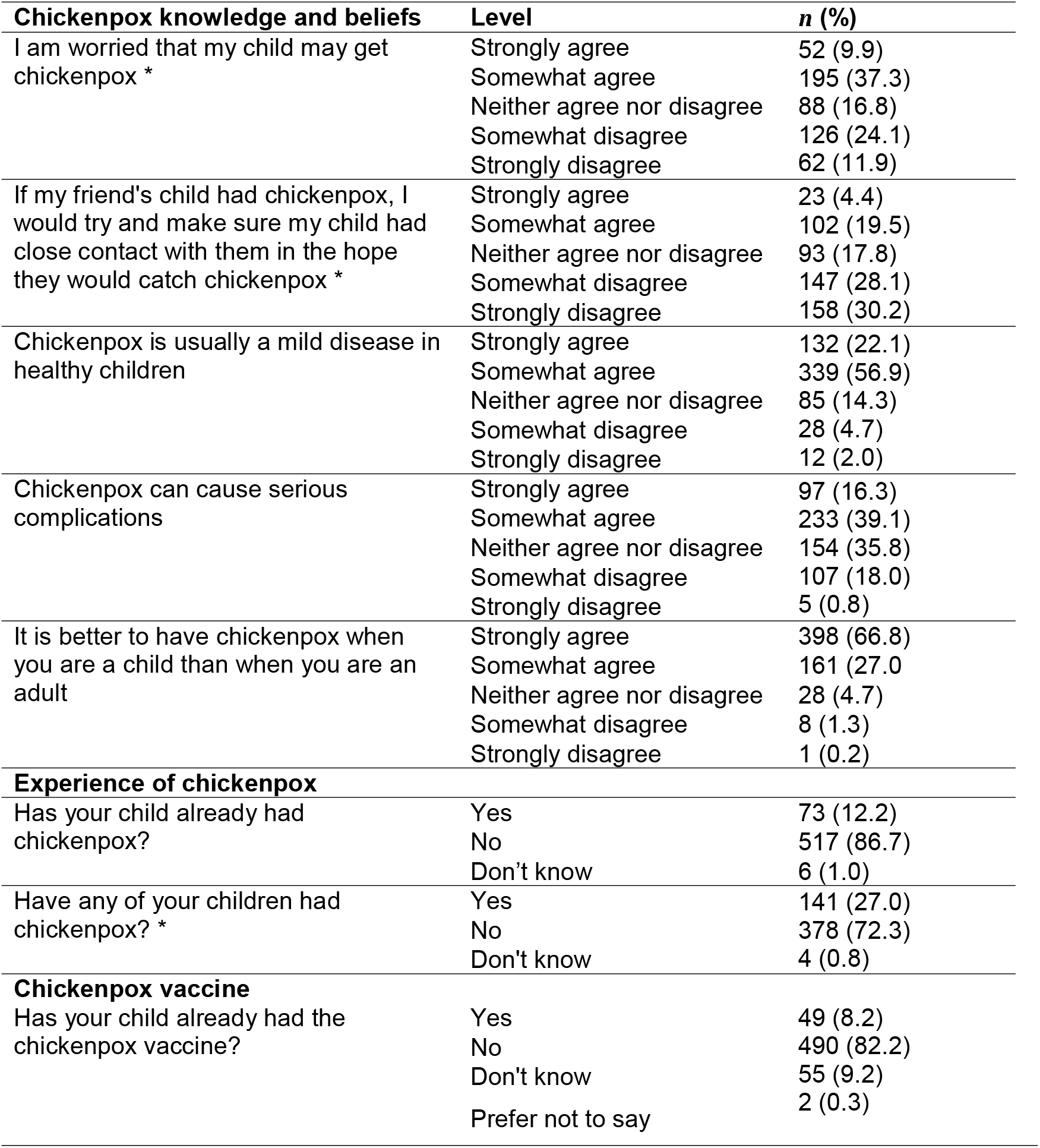
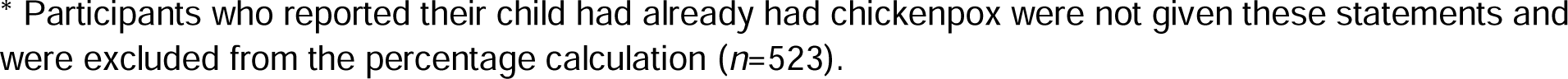
Parents’ attitudes to and experience of chickenpox disease

| **Chickenpox knowledge and beliefs** | **Level** | **n (%)** |
| --- | --- | --- |
| I am worried that my child may get chickenpox * | Strongly agree | 52 (9.9) |
|  | Somewhat agree | 195 (37.3) |
|  | Neither agree nor disagree | 88 (16.8) |
|  | Somewhat disagree | 126 (24.1) |
|  | Strongly disagree | 62 (11.9) |
| If my friend's child had chickenpox, I would try and make sure my child had close contact with them in the hope they would catch chickenpox * | Strongly agree | 23 (4.4) |
|  | Somewhat agree | 102 (19.5) |
|  | Neither agree nor disagree | 93 (17.8) |
|  | Somewhat disagree | 147 (28.1) |
|  | Strongly disagree | 158 (30.2) |
| Chickenpox is usually a mild disease in healthy children | Strongly agree | 132 (22.1) |
|  | Somewhat agree | 339 (56.9) |
|  | Neither agree nor disagree | 85 (14.3) |
|  | Somewhat disagree | 28 (4.7) |
|  | Strongly disagree | 12 (2.0) |
| Chickenpox can cause serious complications | Strongly agree | 97 (16.3) |
|  | Somewhat agree | 233 (39.1) |
|  | Neither agree nor disagree | 154 (35.8) |
|  | Somewhat disagree | 107 (18.0) |
|  | Strongly disagree | 5 (0.8) |
| It is better to have chickenpox when you are a child than when you are an adult | Strongly agree | 398 (66.8) |
|  | Somewhat agree | 161 (27.0) |
|  | Neither agree nor disagree | 28 (4.7) |
|  | Somewhat disagree | 8 (1.3) |
|  | Strongly disagree | 1 (0.2) |
| **Experience of chickenpox** |  |  |
| Has your child already had chickenpox? | Yes | 73 (12.2) |
|  | No | 517 (86.7) |
|  | Don't know | 6 (1.0) |
| Have any of your children had chickenpox? * | Yes | 141 (27.0) |
|  | No | 378 (72.3) |
|  | Don't know | 4 (0.8) |
| **Chickenpox vaccine** |  |  |
| Has your child already had the chickenpox vaccine? | Yes | 49 (8.2) |
|  | No | 490 (82.2) |
|  | Don't know | 55 (9.2) |
|  | Prefer not to say | 2 (0.3) |

\* Participants who reported their child had already had chickenpox were not given these statements and were excluded from the percentage calculation ( $n=523$ ).

**Table 3.** Parental intention to vaccinate; *n* (%).

| | | Before<br>information<br>($n=547$) * | After<br>information<br>($n=546$) * |
| --- | --- | --- | --- |
| If a vaccine to protect against chickenpox was recommended in the UK, how likely is it that you would accept it for your child? | Extremely likely | 231 (42.2) | 233 (42.6) |
|  | Somewhat likely | 185 (33.8) | 171 (31.3) |
|  | Neither likely nor unlikely | 42 (7.7) | 42 (7.7) |
|  | Somewhat unlikely | 53 (9.7) | 62 (11.3) |
|  | Extremely unlikely | 36 (6.6) | 38 (6.9) |

\* Participants whose children had already had chickenpox vaccine were not asked how likely they would accept a chickenpox vaccine and they are not included in the totals.

**Table 4.** Parental intention to accept different options for administration of the varicella vaccine (*n* = 596)

| Item | Level | *n* (%) |
| --- | --- | --- |
| **First dose of chickenpox vaccine** |  |  |
| First dose of chickenpox vaccine given in combination with MMR vaccine as a 4-in-1 vaccine (MMRV) | Extremely likely | 187 (31.4) |
|  | Somewhat likely | 204 (34.2) |
|  | Neither likely nor unlikely | 51 (8.6) |
|  | Somewhat unlikely | 89 (14.9) |
|  | Extremely unlikely | 65 (10.9) |
| First dose as a separate vaccine at the same time as the vaccines given at 12 months | Extremely likely | 104 (17.5) |
|  | Somewhat likely | 173 (29.0) |
|  | Neither likely nor unlikely | 77 (12.9) |
|  | Somewhat unlikely | 142 (23.8) |
|  | Extremely unlikely | 99 (16.6) |
|  | Left blank | 1 (0.2) |
| First dose of chickenpox vaccine given separately at another visit after the vaccines given at 12 months | Extremely likely | 169 (28.4) |
|  | Somewhat likely | 198 (33.2) |
|  | Neither likely nor unlikely | 74 (12.4) |
|  | Somewhat unlikely | 96 (16.1) |
|  | Extremely unlikely | 59 (9.9) |
| **Second dose of chickenpox vaccine** |  |  |
| Second dose of chickenpox vaccine given separately at another visit at 15-18 months | Extremely likely | 172 (28.9) |
|  | Somewhat likely | 208 (34.9) |
|  | Neither likely nor unlikely | 65 (10.9) |
|  | Somewhat unlikely | 99 (16.6) |
|  | Extremely unlikely | 52 (8.7) |
| Second dose of chickenpox vaccine given in combination with the pre-school dose of MMR vaccine as a 4-in-1 vaccine (MMRV) at 3 years 4 months | Extremely likely | 187 (31.4) |
|  | Somewhat likely | 220 (36.9) |
|  | Neither likely nor unlikely | 59 (9.9) |
|  | Somewhat unlikely | 74 (12.4) |
|  | Extremely unlikely | 56 (9.4) |
| Second dose of chickenpox vaccine given as a separate vaccine at the same time as the pre-school MMR vaccine | Extremely likely | 119 (20.0) |
|  | Somewhat likely | 200 (33.6) |
|  | Neither likely nor unlikely | 90 (15.1) |
|  | Somewhat unlikely | 117 (19.6) |
|  | Extremely unlikely | 70 (11.7) |
| **Injections per visit** |  |  |
| How many injections would you want your child to have on any one occasion? | No more than 2 | 183 (30.7) |
|  | No more than 3 | 61 (10.2) |
|  | No more than 4 | 93 (15.6) |
|  | No more than 5 | 5 (0.8) |
|  | I would be happy to go along with whatever the NHS recommends | 241 (40.4) |
|  | Not applicable | 13 (2.2) |

### Effect of information on parents’ acceptance of a varicella vaccine

Before reading information on varicella infection and vaccine, 76.0% of participants responded that they would be extremely or somewhat likely to accept a varicella vaccine for their child; this changed little after information about the disease and vaccine was provided, with 73.9% extremely or somewhat likely to accept the vaccine (Table 3). Respondents were asked to describe the main reason they would be likely or unlikely to accept a varicella vaccine. The content analysis generated 115 unique codes. The most frequent codes generated from those participants who were likely to accept the vaccine were Protect child (*n*=158), Avoid complications (*n*=58) and Follow recommendations (*n*=38). The most frequent codes from those who were uncertain, were Need more info (*n*=8), Mild illness (*n*=6), Already had CP (*n*=3). The most frequent codes for those who were unlikely, were Mild illness (*n*=30), Unnecessary (*n*=20), Chickenpox complications rare (*n*=10). The full list of codes is in the Supplementary Materials.

### Attitudes towards options for administration of varicella vaccine

Two thirds of respondents (65.6%) reported they would be extremely or somewhat likely to accept a varicella vaccine for their child if it were given as a combination vaccine at 12 months of age at the same time as other vaccines. Fewer parents (46.5%) would be extremely or somewhat likely to accept the vaccine at 12 months if it were given separately with the other vaccines. If the vaccine was offered separately at another vaccine visit, 61.6% reported they would be extremely or somewhat likely to accept it (Table 4). Responses to items in the vaccine hesitancy scale are shown in Table 5. 93.6% of parents indicated that they had never refused a vaccine for their child/children, with only 5.9% indicating that they had.

**Table 5.** General vaccination views (*n* = 596)

| Item | Level | *n* (%) |
| --- | --- | --- |
| **Vaccine hesitancy scale** |  |  |
| Childhood vaccines are important for my child/children's health* | Strongly agree | 451 (75.7) |
|  | Somewhat agree | 123 (20.6) |
|  | Neither agree nor disagree | 10 (1.7) |
|  | Somewhat disagree | 7 (1.2) |
|  | Strongly disagree | 5 (0.8) |
| Childhood vaccines are effective ('effective' means that most vaccines give good protection)* | Strongly agree | 430 (72.1) |
|  | Somewhat agree | 136 (22.8) |
|  | Neither agree nor disagree | 20 (3.4) |
|  | Somewhat disagree | 5 (0.8) |
|  | Strongly disagree | 5 (0.8) |
| Vaccines are generally safe ('safe' means serious side effects are rare)* | Strongly agree | 371 (62.2) |
|  | Somewhat agree | 175 (29.4) |
|  | Neither agree nor disagree | 36 (6.0) |
|  | Somewhat disagree | 8 (1.3) |
|  | Strongly disagree | 6 (1.0) |
| Having my child/children vaccinated is important for the health of others in my community* | Strongly agree | 395 (66.3) |
|  | Somewhat agree | 139 (23.3) |
|  | Neither agree nor disagree | 38 (6.4) |
|  | Somewhat disagree | 13 (2.2) |
|  | Strongly disagree | 11 (1.8) |
| All childhood vaccines offered by the NHS are beneficial* | Strongly agree | 384 (64.4) |
|  | Somewhat agree | 139 (23.3) |
|  | Neither agree nor disagree | 50 (8.4) |
|  | Somewhat disagree | 15 (2.5) |
|  | Strongly disagree | 8 (1.3) |
| New vaccines carry more risk than older vaccines<sup>#</sup> | Strongly agree | 56 (9.4) |
|  | Somewhat agree | 153 (25.7) |
|  | Neither agree nor disagree | 173 (29.0) |
|  | Somewhat disagree | 143 (24.0) |
|  | Strongly disagree | 71 (11.9) |
| The information I receive about vaccines from the NHS is reliable* | Strongly agree | 315 (52.9) |
|  | Somewhat agree | 197 (33.1) |
|  | Neither agree nor disagree | 62 (10.4) |
|  | Somewhat disagree | 14 (2.3) |
|  | Strongly disagree | 8 (1.3) |
| Getting vaccines is a good way to protect my child/children from disease* | Strongly agree | 422 (70.8) |
|  | Somewhat agree | 143 (24.0) |
|  | Neither agree nor disagree | 18 (3.0) |
|  | Somewhat disagree | 8 (1.3) |
|  | Strongly disagree | 5 (0.8) |
| Generally I do what my doctor or health care provider recommends about vaccines for my child/children* | Strongly agree | 345 (57.9) |
|  | Somewhat agree | 171 (28.7) |
|  | Neither agree nor disagree | 54 (9.1) |
|  | Somewhat disagree | 21 (3.5) |
|  | Strongly disagree | 5 (0.8) |
| I am concerned about serious adverse effects of vaccines<sup>#</sup> | Strongly agree | 60 (10.1) |
|  | Somewhat agree | 192 (32.2) |
|  | Neither agree nor disagree | 130 (21.8) |
|  | Somewhat disagree | 162 (27.2) |
|  | Strongly disagree | 52 (8.7) |

\* Items comprising the 'lack of confidence' hesitancy subscale (scored 1 = strongly agree, 5 = strongly disagree). <sup>#</sup> Items comprising the 'risk' hesitancy subscale (scored 1 = strongly disagree, 5 = strongly agree).

The sources from which parents get their information about vaccinations from are detailed in Table 6, including their main source of information. A breakdown of specific internet sites used is provided in Supplementary Materials. Table 7 details the level of trust parents have in different information sources.

**Table 6.** Sources of vaccination information (*n* = 596)

| **General sources** |  |  |
| --- | --- | --- |
| **Item** | **Source of vaccination information *n* (%)** | **Main source of vaccination information *n* (%)** |
| Your GP | 541 (90.8) | 207 (34.7) |
| Practice nurse/Nurse in GP clinic | 336 (56.4) | 91 (15.3) |
| Health Visitor | 382 (64.1) | 78 (13.1) |
| Midwife | 189 (31.7) | 3 (0.5) |
| Pharmacist | 56 (9.4) | 1 (0.2) |
| School nurse | 58 (9.7) | 0 (0) |
| NHS.uk | 374 (62.8) | 110 (18.5) |
| Alternative health practitioner e.g. homeopath | 7 (1.2) | 0 (0) |
| NHS leaflets | 209 (35.1) | 15 (2.5) |
| NHS 111 telephone service | 28 (4.7) | 0 (0) |
| Red book/Personal Child Health Record | 270 (45.3) | 29 (4.9) |
| Other parents | 122 (20.5) | 0 (0) |
| Friends | 122 (20.5) | 3 (0.5) |
| Family | 156 (26.2) | 3 (0.5) |
| Television | 31 (5.2) | 0 (0) |
| Books | 25 (4.2) | 3 (0.5) |
| Newspapers | 22 (3.7) | 0 (0) |
| Magazines | 11 (1.8) | 0 (0) |
| Internet | 219 (36.7) | 48 (8.1) |
| Not applicable | 1 (0.2) | 1 (0.2) |
| Other (please describe) | 6 (1.0) | 2 (0.3) |
| Left blank | 0 (0.0) | 2 (0.3) |

**Table 7.**
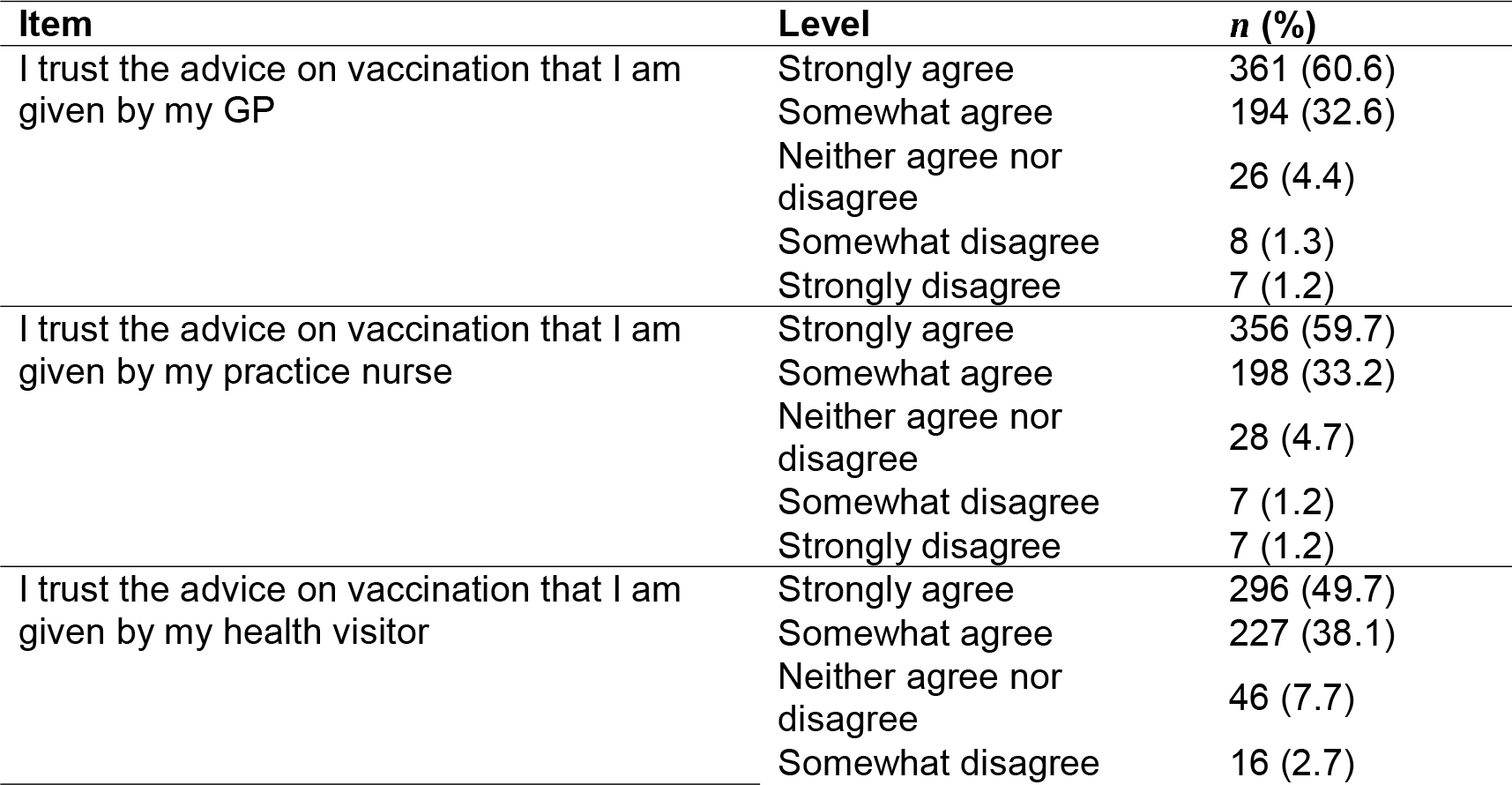
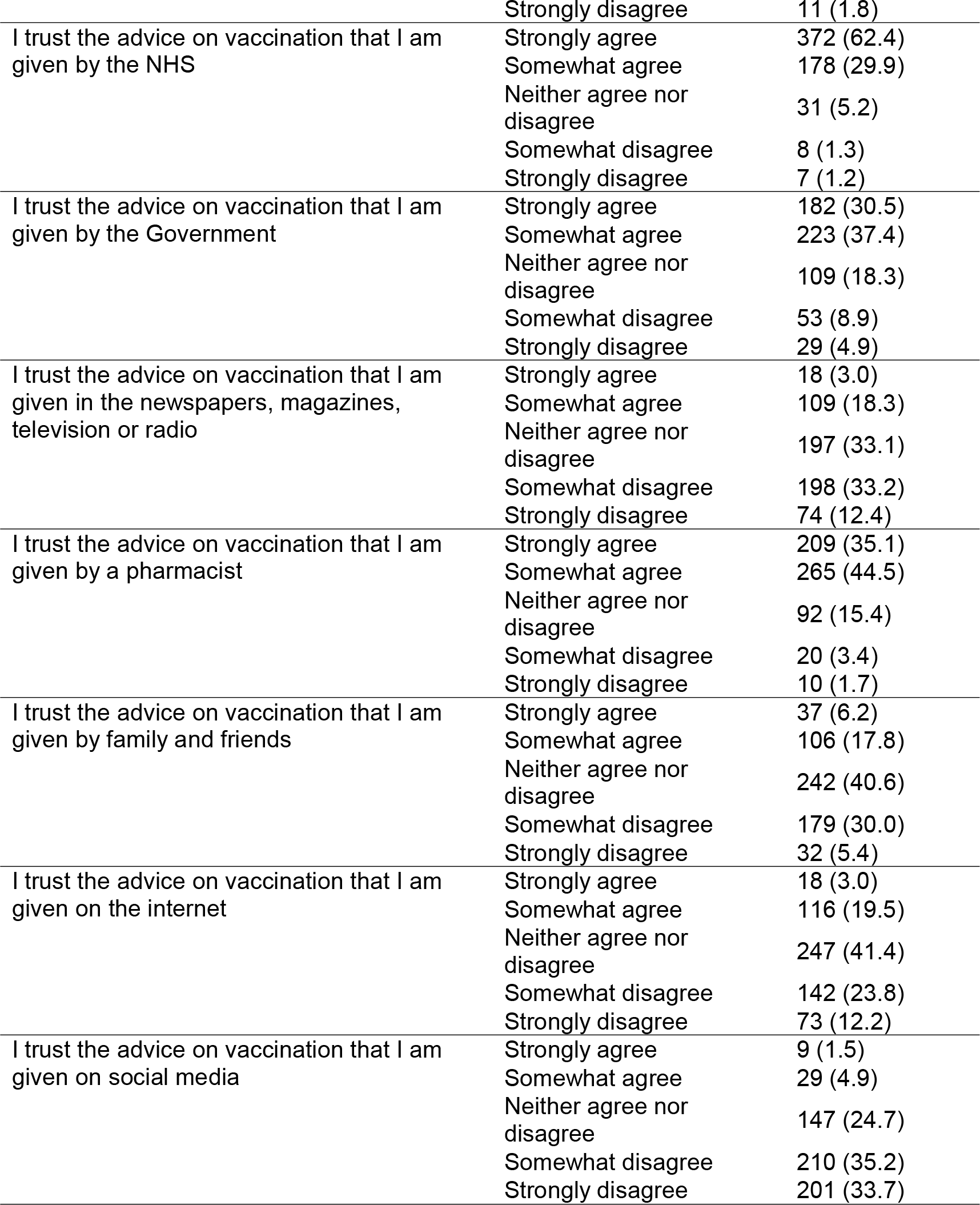
Trust in vaccination sources (*n* = 596)

Based on the percentage of those somewhat or strongly agreeing with the statement “I trust advice” from each source, the most trusted sources were reported to be GPs (93.2%), practice nurses (92.9%), the NHS (92.3%), health visitors (87.8%), pharmacists (79.6%) and the government (67.9%) (Table 7). The least trusted source was social media (6.4% somewhat or strongly agreeing).

In our regression analysis (Table 8), parental age and gender were non-significant predictors of vaccination intention; the other predictors were significant. For the vaccination hesitancy subscales, higher scores on ‘lack of confidence’ and ‘risk’ indicated lower odds (less likelihood) of a higher score on the intention scale. Across the categories of the ‘chicken pox is a mild disease in healthy children’ variable, there was a weak (*R*^2^ change = .016) negative association with vaccination intention, indicating that stronger agreement with this statement predicts weaker vaccination intention. For the ‘chickenpox can cause serious complications’ variable, there was a somewhat stronger (*R*^2^ change = .054) positive association (*d* = .237), indicating that stronger agreement with this statement predicts stronger vaccination intention.

**Table 8.** Results of the ordinal logistic regression analysis. The outcome variable is the likelihood of vaccination (five-point scale from extremely unlikely to extremely likely). The association between an ordinal predictor and the outcome variable is quantified by the change in the Nagelkerke *R*^2^ when the predictor is added to the model. *n* = 536. Nagelkerke *R*^2^ for full model is .481

| | Odds ratio* | 95% Wald confidence interval | Change in Nagelkerke $R^2$ | $p$ value |
| --- | --- | --- | --- | --- |
| Age | 0.983 | 0.950, 1.017 |  | .331 |
| Gender (reference: male) | 0.838 | 0.548, 1.281 |  | .414 |
| Refused vaccine for children (reference: yes) | 0.440 | 0.204, 0.949 |  | .036 |
| Index child has already had chickenpox (reference: yes) | 0.419 | 0.247, 0.710 |  | .001 |
| Chickenpox is usually a mild disease in healthy children (reference: strongly agree) |  |  | .016 | .011 |
| Strongly disagree | 1.575 | 0.334, 7.430 |  |  |
| Somewhat disagree | 3.316 | 1.253, 8.775 |  |  |
| Neither | 2.800 | 1.490, 5.261 |  |  |
| Somewhat agree | 1.538 | 0.997, 2.373 |  |  |
| Chickenpox can cause serious complications (reference: strongly agree) |  |  | .054 | <.001 |
| Strongly disagree | 0.071 | 0.008, 0.620 |  |  |
| Somewhat disagree | 0.211 | 0.111, 0.403 |  |  |
| Neither | 0.229 | 0.125, 0.421 |  |  |
| Somewhat agree | 0.612 | 0.345, 1.084 |  |  |
| Vaccine hesitancy: |  |  |  |  |
| Lack of confidence (8–40) | 0.792 | 0.755, 0.831 |  | <.001 |
| Risk (2–10) | 0.774 | 0.695, 0.861 |  | <.001 |

\* The odds ratios indicate the increase or decrease in the odds of a one-unit increase in the outcome variable for a one-unit increase in the predictor variable for continuous predictors, or for a shift from the reference category to the category in question for binary predictors. For an ordinal predictor the odds ratio is cumulative; it indicates the increase or decrease in the odds of a one-unit increase in the outcome variable for being a particular category in the predictor variable compared with being in any one of the lower categories.

## Discussion

We found high levels of acceptability of a varicella vaccine among UK parents of children under five years of age, with preference for a combined MMRV vaccine or varicella vaccination delivered at an additional immunisation visit rather than as an additional vaccination at an existing visit. Higher likelihood of accepting the vaccine was associated with agreeing that varicella can cause complications, while lower likelihood of acceptance was associated with agreeing that varicella is normally a mild illness and also with a lack of confidence in vaccines and concern about risk (generally and of serious adverse events) as measured by the vaccine hesitancy scale. Our content analysis provides more detail, with parents who were likely to accept the vaccine being most concerned with protecting their child (from complications, from suffering in adulthood and from experiencing the discomfort of chickenpox) and other people, especially vulnerable others.

Our findings provide a snapshot of views in the midst of the COVID-19 pandemic when a much-publicised COVID-19 vaccine programme was being rolled out for adults. While this may have affected responses, making parents more aware of vaccination (both positively and negatively), the positive attitudes to a childhood vaccination reported by our participants reflect other UK studies of attitudes towards childhood vaccines generally [16] and attitudes towards specific vaccines, including new ones such as the meningococcal group B (MenB) vaccine.[18] Our study confirms that parents value vaccination, consider it to be important, and are receptive to the inclusion of a varicella vaccine in the routine programme. Although they have a preference for their child having fewer injections, many parents would still accept a varicella vaccine even if this required an additional injection, with a clear demonstration of trust in health care professionals and in the NHS and a willingness to follow its recommendations.

To our knowledge, this is the largest recent study of parental views about varicella conducted in the UK among parents of young children. The sample is broadly representative for ethnicity and there was a high completion rate. One possible limitation is that despite it being a common childhood illness, only 12% of youngest children were reported to have had chickenpox and only 27% of those whose youngest children had not had it reported that their other children had. This may have been affected by the timing of the survey, which took place during the COVID-19 pandemic: a time when reports of other childhood infections such as measles have also been much less common due to public health measures such as lockdowns, social distancing and restrictions on overseas travel.[19] It is possible this lack of direct experience of varicella may have influenced parents’ responses. A second limitation is that our sample is unlikely to capture the views of parents who are vaccine hesitant or who are likely to refuse vaccines for their children, with 93.6% of our sample reporting never having refused a vaccine for their child. A third limitation is that although the sample was broadly representative for ethnicity, the sample was generally more educated and with a higher income than the population of the UK, and since the data were collected online, only parents with internet access were represented.

The WHO recommends introducing routine varicella vaccination only if 80% or greater coverage can be achieved.[20] Although our study suggests that the majority of parents would be in favour of having their child vaccinated against varicella, the well documented intention-behaviour gap,[21] whereby intention is usually higher than behaviour, means that steps would need to be taken to maximise uptake. Delivering the vaccine in line with parental preferences would be one such step and our data suggest that this should involve a combined MMRV vaccination, which would serve to avoid an increase in needle burden or in the number of appointments needed. It is also somewhat reassuring that more than 80% of parents in our survey report usually doing following the advice of their healthcare provider.

Although there was little evidence of vaccine hesitancy in our cohort as measured by the vaccine hesitancy scale, responses were uneven to the question about whether new vaccines carry more risk that old vaccines. It will be important to emphasise to parents that the varicella vaccination is well established, having been administered routinely in many countries for many years and been available privately in the UK. Another implication of our findings for vaccine communication is a need to advise parents that although chickenpox is usually a mild childhood disease, it can have serious side effects. In our open-ended question, for those parents who reported being unlikely to have the vaccine for their child, one of the most frequently reported reasons was that chickenpox is a mild disease and this was also a predictor of lower likelihood of acceptance in our regression analysis.

Our study has provided some insight into the likely views and preferences of parents should varicella be added to the routine childhood immunisation schedule in the UK. It would be useful to also understand knowledge and views of health care professionals involved in vaccine programmes who would be recommending and administering the vaccine, and research is underway to capture this information.

## Conclusion

Our survey of UK parents reveals that introducing varicella vaccination to the routine childhood immunisation schedule would likely be well received. By introducing it as a combined MMRV vaccination, and by advising parents that chickenpox can have complications and emphasising that varicella is a well-established and safe vaccine, there is the real potential to achieve WHO-recommended levels of uptake and significantly reduce the burden of both serious complications and unpleasant albeit mild symptoms from this common childhood infection on families and the NHS.

## Data Availability

Data will be made available online on acceptance of manuscript.

## Acknowledgements

We would like to thank Elizabeth Kaplunov for comments on an earlier version of this manuscript.

## Transparency declaration

The authors affirm that the manuscript is an honest, accurate, and transparent account of the study being reported; that no important aspects of the study have been omitted; and that any discrepancies from the study as originally planned have been explained.

## Data sharing statement

Data are available online (LINK TO FOLLOW).

## Appendix

Information provided to all survey participants about varicella and varicella vaccination.

### Chickenpox

Chickenpox is caused by a virus which spreads very easily. Most people have the infection in early childhood when it is usually a mild disease. Most children with chickenpox have a fever, an itchy rash and are unwell for a few days. Serious complications can occur but are rare, these include pneumonia and other serious infections. Chickenpox is more serious in adults and in children who have a problem fighting infections.

### Chickenpox vaccine

The chickenpox vaccine is given from the age of 12 months. It is given by injection. In the UK, two doses of the vaccine are recommended with a gap of at least 4 weeks between the doses. Chickenpox vaccine can be given as a separate vaccine at the same time as other vaccines, or as a combined 4-in-1 vaccine with measles, mumps and rubella (MMRV). Both the separate and combined vaccines are safe with side effects including tenderness where the injection was given and occasionally a mild fever. However, for children under two years of age, there is a very small increased risk of a child having a fever fit when they are given the combination MMRV vaccine compared with the MMR vaccine and a separate chickenpox vaccine given on the same day.

## Supplementary Materials

**Supplementary Table 1.** Most frequent codes generated by content analysis of reasons for likelihood of having, or not having, the varicella vaccination, by likelihood of having the vaccination (likely, uncertain, likely). Codes are presented in descending order of frequency in each category.

|  | **Likely** |  | **Uncertain** |  | **Unlikely** |
| --- | --- | --- | --- | --- | --- |
| Protect child | 158 | Need more information | 8 | Mild illness | 30 |
| Avoid complications | 58 | Mild illness | 6 | Unnecessary | 20 |
| Follow recommendations | 38 | Already had CP | 3 | CP complications rare | 10 |
| Protect (benefits outweigh risks) | 35 | Concern re side effects | 3 | Prefer child to catch virus | 8 |
| Protect (discomfort) | 31 | Personal experience (mild) | 3 | Better to catch virus for immune system | 7 |
| Personal experience (negative) | 28 | Undecided | 3 | Child is healthy | 7 |
| Protect others | 25 | If necessary to protect child | 2 | Risks outweigh benefits | 7 |
| Protect during adulthood | 21 | Something we all go through | 2 | Concern re side effects | 6 |
| Believe in vaccines | 16 | Unnecessary | 2 | Already had it | 5 |
| Protect against scarring | 10 | Based on research about vaccine | 1 | Concern re fever fit | 5 |
| Reduce severity | 10 | Concern re safety | 1 | Effective for limited time | 5 |
| Boost immune system | 9 | Controversy re vaccines | 1 | Prefer to catch as child | 5 |

**Supplementary Table 2.**
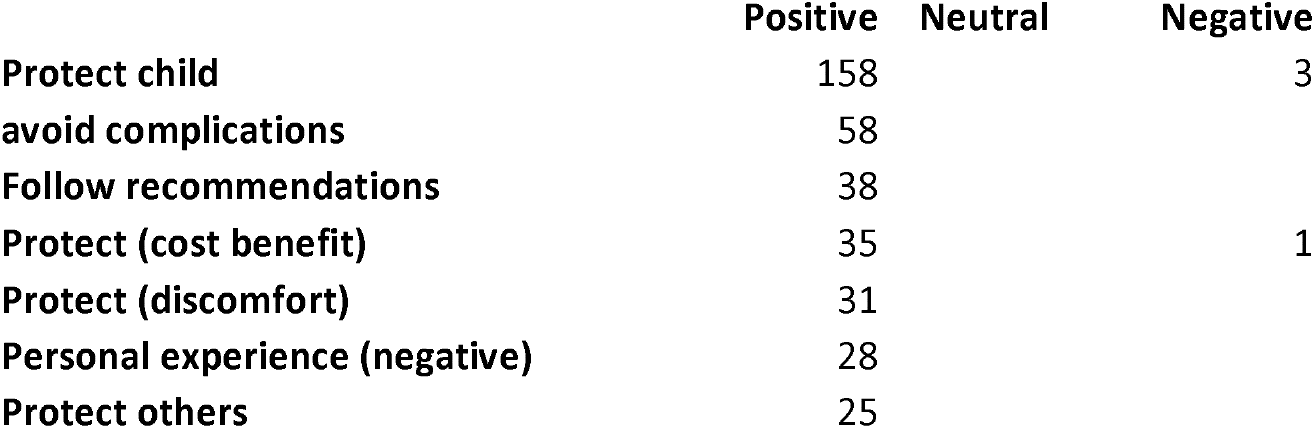
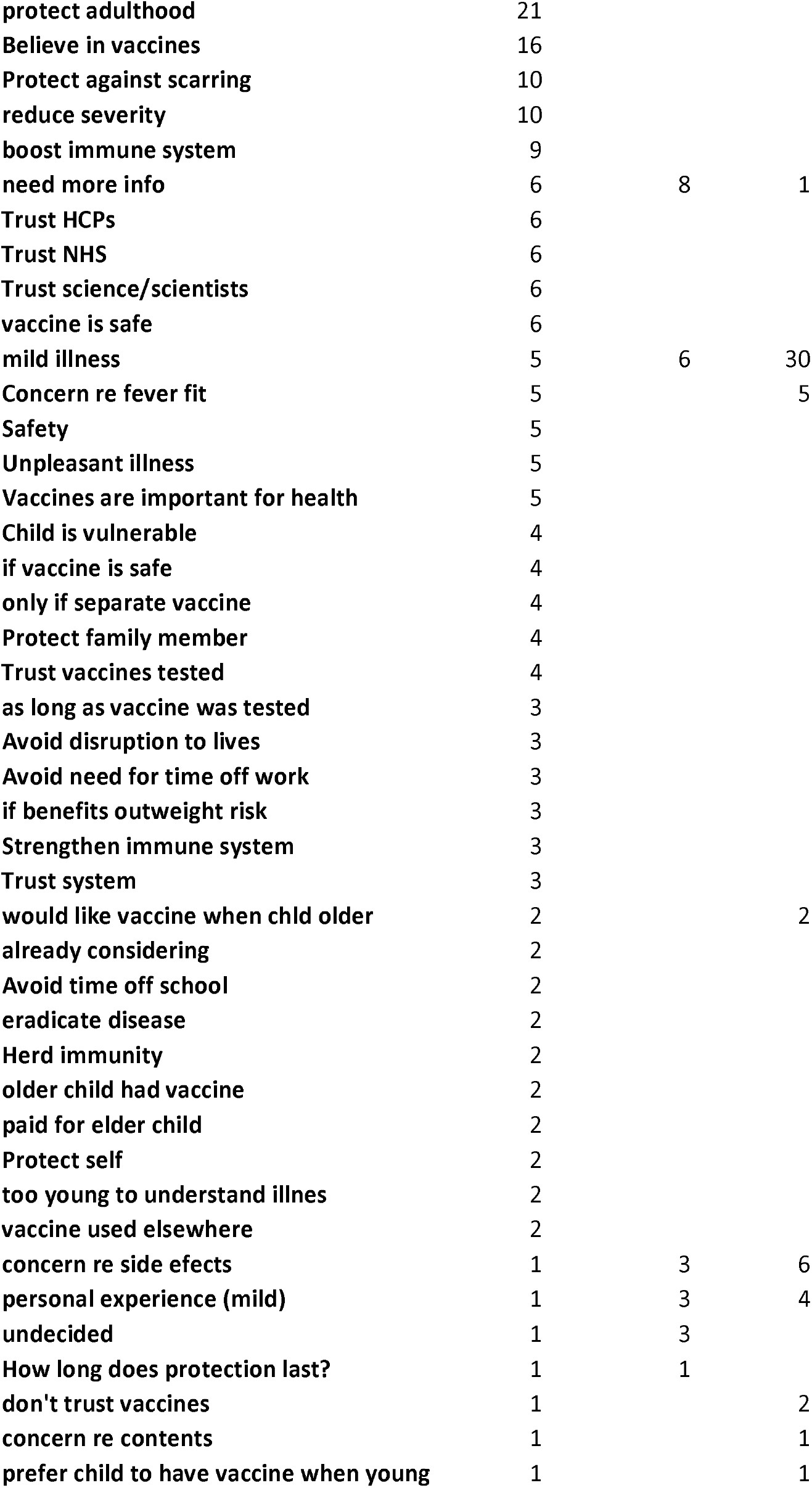
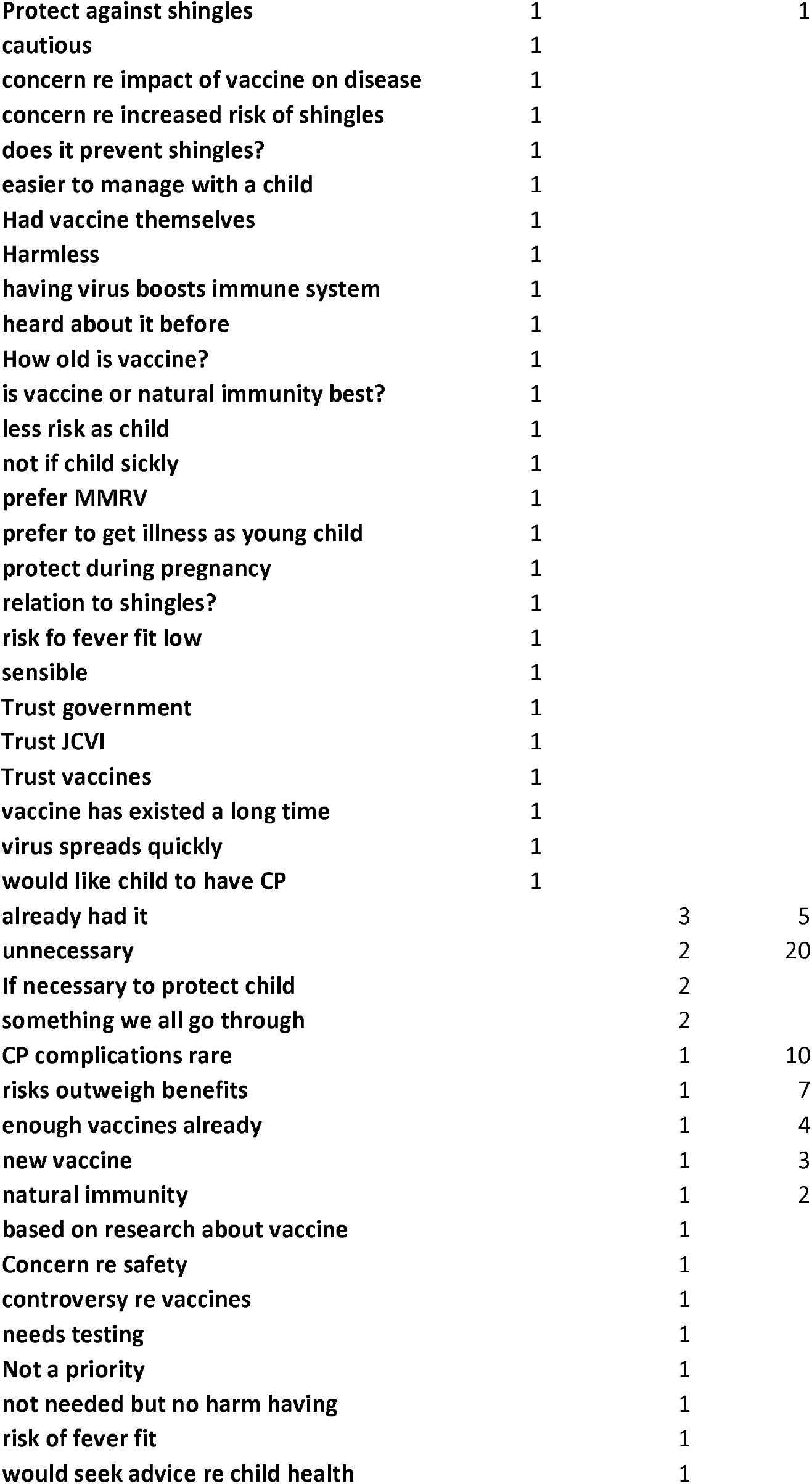
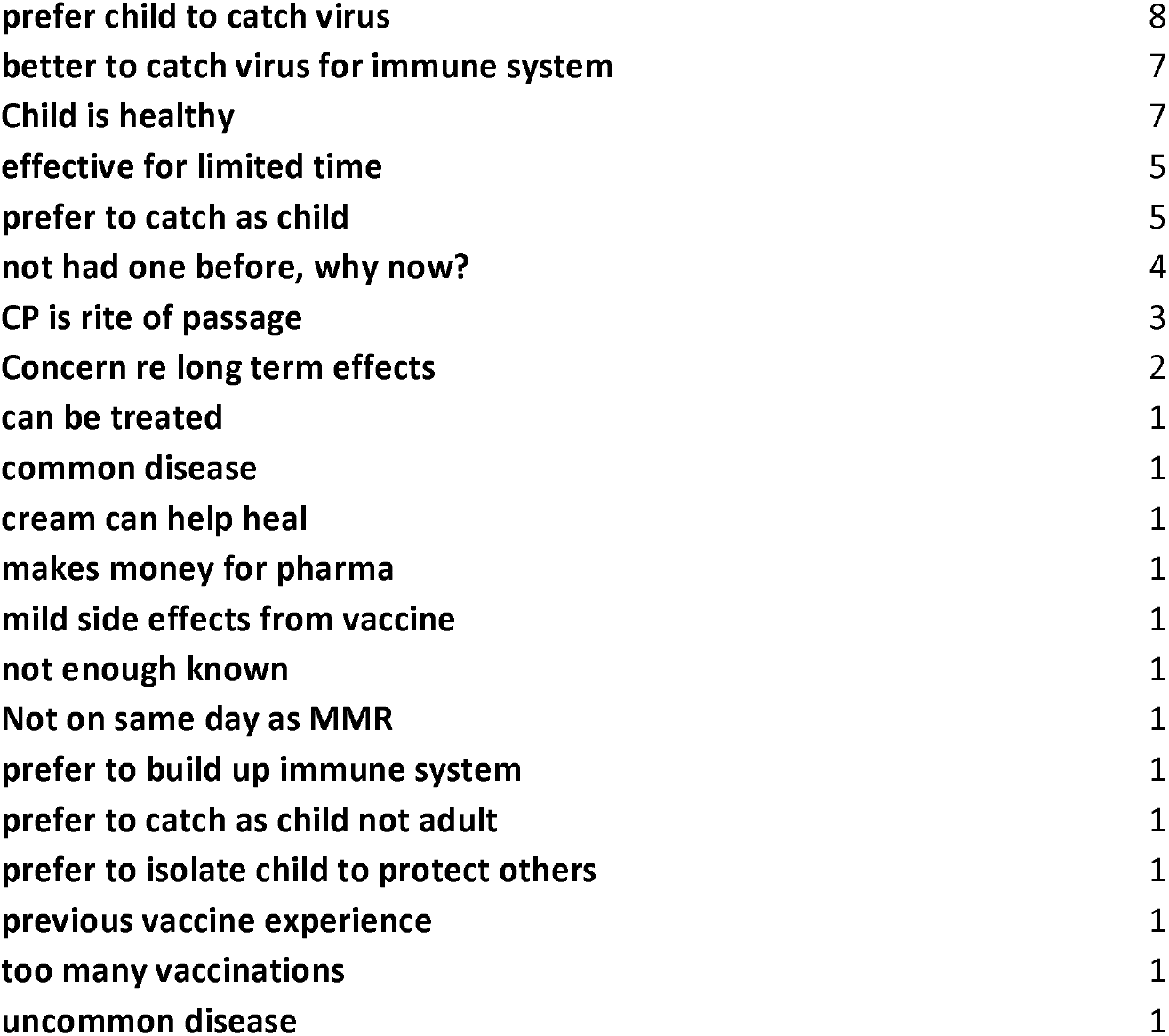
Codes generated by the content analysis of why participants were likely or unlikely to accept the varicella vaccination for their child.

**Supplementary Table 3.** Details of which internet sites parents reported using to find out more about vaccinations.

| **Internet** |  |  |
| --- | --- | --- |
| **Item** | **Level** | **n (%)** |
| Have you ever used the internet to find out more about vaccinations? # | 1 (Yes) | 264 (70.0) |
|  | 2 (No) | 111 (29.4) |
|  | 3 (Don't know) | 2 (5.3) |
| What websites have you used to find out more about vaccinations? * | GOV.UK | 212 (43.7) |
|  | Mumsnet | 91 (18.8) |
|  | Patient.co.uk | 30 (6.2) |
|  | NHS.uk | 429 (88.5) |
|  | Net doctor | 18 (3.7) |
|  | Facebook or Twitter | 35 (7.2) |
|  | Chat rooms or discussion forums | 32 (6.6) |
|  | netmums | 37 (7.6) |
|  | Other (please specify) | 24 (4.9) |
|  | Don't know | 12 (2.5) |

## References

1. Manikkavasagan G, Dezateux C, Wade A, Bedford H. The epidemiology of chickenpox in UK 5-year olds: an analysis to inform vaccine policy. Vaccine. 2010 Nov 10;28(48):7699–705.

2. Cameron JC, Allan G, Johnston F, Finn A, Heath PT, Booy R. Severe complications of chickenpox in hospitalised children in the UK and Ireland. Arch Dis Child 2007;92(12):1062–1066.

3. UK Health Security Agency (2019). Varicella: the green book, chapter 34. Available at: https://www.gov.uk/government/publications/varicella-the-green-book-chapter-34. (Accessed 5th July 2022).

4. Bernal JL, Hobbelen P, Amirthalingam G. Burden of varicella complications in secondary care, England, 2004 to 2017. Euro\ Surveill 2019;24(42):1900233. https://doi.org/10.2807/1560-7917.ES.2019.24.42.1900233

5. Hobbelen PH, Stowe J, Amirthalingam G, Miller L, van Hoek AJ. The burden of hospitalisation for varicella and herpes zoster in England from 2004 to 2013. Journal of Infection. 2016 Sep 1;73(3):241–53.

6. Marin M, Marti M, Kambhampati A, Jeram SM, Seward JF. Global varicella vaccine effectiveness: a meta-analysis. Pediatrics 2016;137(3):e20153741.

7. Klein NP, Fireman B, Yih WK, et al. Measles-mumps-rubella-varicella combination vaccine and the risk of febrile seizures. Pediatrics 2010;126(1):e1–e8.

8. Wutzler P, Bonanni P, Burgess M, Gershon G, Sáfadi MA, Giacomo Casabona G. Varicella vaccination – the global experience, Expert Rev Vaccines 2017;16:8, 833-843, DOI: 10.1080/14760584.2017.1343669

9. UK Health Security Agency (2020). Immunisation against infectious disease. Available at: https://www.gov.uk/government/collections/immunisation-against-infectious-disease-the-green-book#the-green-book. (Accessed 30th March 2022).

10. Joint Committee on Vaccination and Immunisation Varicella Sub-Committee. (2015). Minute of the meeting on 19 June 2015. England. https://app.box.com/s/vdlafy8wm4t5asq2qyfpc4dw6fzeyypf/file/149454406336 (Accessed 17/06/2022).

11. Helmuth IG, Poulsen A, Suppli CH, Mølbak K. Varicella in Europe—a review of the epidemiology and experience with vaccination. Vaccine 2015;33(21):2406–2413.

12. Bedford H, Lansley M. More vaccines for children? Parents’ views. Vaccine. 2007 Nov 7;25(45):7818–23.

13. Lee E, Turner J, Bate J. Parental opinions on childhood varicella and the varicella vaccine: a UK multicentre qualitative interview study. Arch Dis Child 2011;96(9):901. https://doi.org/10.1136/archdischild-2011-300165

14. Larson HJ, Jarrett C, Schulz WS, Chaudhuri M, Zhou Y, Dube E, Schuster M, MacDonald NE, Wilson R. Measuring vaccine hesitancy: the development of a survey tool. Vaccine 2015;33(34):4165–4175. https://doi.org/10.1016/j.vaccine.2015.04.037

15. Shapiro GK, Tatar O, Dube E, Amsel R, Knauper B, Naz A, Perez S, Rosberger Z. The vaccine hesitancy scale: Psychometric properties and validation. Vaccine 2018;36(5):660– 667. https://doi.org/10.1016/j.vaccine.2017.12.043

16. Campbell H, Edwards A, Letley L, Bedford H, Ramsay M, Yarwood J. Changing attitudes to childhood immunisation in English parents. Vaccine 2017;35(22):2979–2985. https://doi.org/10.1016/j.vaccine.2017.03.089

17. Stemler S. An overview of content analysis. Pract Assess Res Eval 2000;7:17. https://doi.org/10.7275/z6fm-2e34

18. Jackson C, Yarwood J, Saliba V, Bedford H. UK parents’ attitudes towards meningococcal group B (MenB) vaccination: a qualitative analysis. BMJ Open 2017;7(4):e012851.

19. UK Health Security Agency (2022). Laboratory confirmed cases of measles, rubella and mums in England: January to March 2022. https://www.gov.uk/government/publications/measles-mumps-and-rubella-lab-confirmed-cases-in-england-2022/laboratory-confirmed-cases-of-measles-rubella-and-mumps-in-england-january-to-march-2022#measles (Accessed 16th June 2022)

20. World Health Organization, 2021. https://cdn.who.int/media/docs/default-source/immunization/immunization_schedules/immunization-routine-table1.pdf?sfvrsn=c7de0e97_9&download=true (Accessed 5th July 20220

21. Sniehotta FF, Scholz U, Schwarzer R. Bridging the intention-behaviour gap: planning, self-efficacy, and action control in the adoption and maintenance of physical exercise. Psychol Health 2005;20(2):143–60. DOI: 10.1080/08870440512331317670

